# Peripheral Transcriptomics in Acute and Long-Term Kidney Dysfunction in SARS-CoV2 Infection

**DOI:** 10.1101/2023.10.25.23297469

**Authors:** Pushkala Jayaraman, Madhumitha Rajagopal, Ishan Paranjpe, Lora Liharska, Mayte Suarez-Farinas, Ryan Thompson, Diane Marie Del Valle, Noam Beckmann, Wonsuk Oh, Faris F. Gulamali, Justin Kauffman, Edgar Gonzalez-Kozlova, Sergio Dellepiane, George Vasquez-Rios, Akhil Vaid, Joy Jiang, Annie Chen, Ankit Sakhuja, Steven Chen, Ephraim Kenigsberg, John Cijiang He, Steven G Coca, Lili Chan, Eric Schadt, Miram Merad, Seunghee Kim-Schulze, Sacha Gnjatic, Ephraim Tsalik, Raymond Langley, Alexander W Charney, Girish N Nadkarni

**Affiliations:** The Charles Bronfman Institute for Personalized Medicine (CBIPM), Icahn School of Medicine at Mount Sinai, New York, NY USA; Samuel Bronfman Department of Medicine The Barbara T Murphy Division of Nephrology Icahn School of Medicine at Mount Sinai, New York, NY USA; Department of Medicine Stanford University, Stanford, CA, USA; Department of Genetics and Genomic Sciences, Icahn School of Medicine at Mount Sinai, New York, NY USA; Department of Population Health Science and Policy Icahn School of Medicine at Mount Sinai, New York, NY USA; Department of Immunology and Immunotherapy Icahn School of Medicine at Mount Sinai, New York, NY USA; Division of Data Driven and Digital Medicine (D3M), Icahn School of Medicine at Mount Sinai, New York, NY USA; The Precision Immunology Institute, Icahn School of Medicine at Mount Sinai, New York, NY USA; Center for Applied Genomics & Precision Medicine Division of Infectious Diseases Duke University School of Medicine Raleigh NC USA; Department of Pharmacology University of South Alabama Mobile AL USA; Department of Psychiatry Icahn School of Medicine at Mount Sinai, New York, NY USA

**Author notes:** **Corresponding Author:** Girish N Nadkarni, MD, MPH Icahn School of Medicine at Mount Sinai, New York, NY, USA.

## Abstract

**Background:** Acute kidney injury (AKI) is common in hospitalized patients with SARS-CoV2 infection despite vaccination and leads to long-term kidney dysfunction. However, peripheral blood molecular signatures in AKI from COVID-19 and their association with long-term kidney dysfunction are yet unexplored.

**Methods:** In patients hospitalized with SARS-CoV2, we performed bulk RNA sequencing using peripheral blood mononuclear cells(PBMCs). We applied linear models accounting for technical and biological variability on RNA-Seq data accounting for false discovery rate (FDR) and compared functional enrichment and pathway results to a historical sepsis-AKI cohort. Finally, we evaluated the association of these signatures with long-term trends in kidney function.

**Results:** Of 283 patients, 106 had AKI. After adjustment for sex, age, mechanical ventilation, and chronic kidney disease (CKD), we identified 2635 significant differential gene expressions at FDR<0.05. Top canonical pathways were *EIF2* signaling, oxidative phosphorylation, mTOR signaling, and Th17 signaling, indicating mitochondrial dysfunction and endoplasmic reticulum (ER) stress. Comparison with sepsis associated AKI showed considerable overlap of key pathways (48.14%). Using follow-up estimated glomerular filtration rate (eGFR) measurements from 115 patients, we identified 164/2635 (6.2%) of the significantly differentiated genes associated with overall decrease in long-term kidney function. The strongest associations were ‘autophagy’, ‘renal impairment via fibrosis’, and ‘cardiac structure and function’.

**Conclusions:** We show that AKI in SARS-CoV2 is a multifactorial process with mitochondrial dysfunction driven by ER stress whereas long-term kidney function decline is associated with cardiac structure and function and immune dysregulation. Functional overlap with sepsis-AKI also highlights common signatures, indicating generalizability in therapeutic approaches.

**SIGNIFICANCE STATEMENT:** Peripheral transcriptomic findings in acute and long-term kidney dysfunction after hospitalization for SARS-CoV2 infection are unclear. We evaluated peripheral blood molecular signatures in AKI from COVID-19 (COVID-AKI) and their association with long-term kidney dysfunction using the largest hospitalized cohort with transcriptomic data. Analysis of 283 hospitalized patients of whom 37% had AKI, highlighted the contribution of mitochondrial dysfunction driven by endoplasmic reticulum stress in the acute stages. Subsequently, long-term kidney function decline exhibits significant associations with markers of cardiac structure and function and immune mediated dysregulation. There were similar biomolecular signatures in other inflammatory states, such as sepsis. This enhances the potential for repurposing and generalizability in therapeutic approaches.

## INTRODUCTION

Acute kidney injury (AKI) is common in hospitalized patients with SARS-CoV2 infection and the clinical syndrome of coronavirus disease-19 (COVID-19).^1,2^ During the COVID-19 pandemic in the United States, AKI incidence was highly variable.^1,3–5^ While the rates of AKI have decreased and even have overall less severity during vaccination, it is still a significant complication.^6,7^ However, the molecular pathophysiology of AKI in COVID-19 is unclear.^2,8^

Previous studies used post-mortem histopathological samples to understand the pathophysiology of COVID-19 associated AKI.^2,9–11^ Although limited by selection bias, acute tubular injury is the most common observation across these studies. Additionally, these studies demonstrated comparable morphological, transcriptomic, and proteomic features between COVID-19 associated AKI (COVID-AKI) and sepsis associated AKI.^12^ However, there have not been any studies on peripheral transcriptomics in general hospitalized populations to complement these findings.

In addition to AKI, COVID-19 is associated with long term kidney function.^13–15^ Decline in kidney function is a major component of post-acute sequelae of SARS-CoV2 (PASC).^16^ However, peripheral transcriptomics linked to long term kidney dysfunction and PASC are unknown, especially if the initial SARS-CoV2 infection required hospitalization.^16^

We have previously shown that acute markers for tubular injury and hemodynamic instability play a role in COVID-AKI and long-term kidney function decline using plasma proteomics.^17^ In this study, we aim to understand how the molecular mechanisms of immune dysfunctions in COVID-AKI are regulated via transcriptomic analysis of peripheral-blood samples, using peripheral blood mononuclear cells (PBMCs) of patients. Specifically, we sought to (1) identify canonical pathways and genes that are differentially expressed in COVID-AKI, (2) understand if AKI in COVID was distinct from other inflammatory states via comparison with a pre-COVID-19 bulk-transcriptomics dataset of sepsis associated AKI, and (3) evaluate whether any identified signatures and/or pathways are associated with long-term kidney dysfunction.

## METHODS

### Patient Cohort

The detailed cohort characteristics and specimen collection procedures are previously described.^18^ For each COVID-19 patient, we defined cases as COVID-19 patients who developed AKI (stage 1, 2 or 3) during their admission (n=106) and controls as all other COVID-19 patients (n=177) without AKI **(Figure 1a)**. We used samples acquired at the last available timepoint during the hospital course **(Figure 1b)**. We excluded patients who developed AKI after the last specimen collection. If a patient had multiple AKI events during their hospitalizations, we included the sample collected after their last AKI event to avoid repeat sampling/confounding.^17^ The Mount Sinai Institutional Review Board approved this study^19^ under a regulatory approval allowing for access to patient-level data and biospecimen collection. This research was reviewed and approved by the Icahn School of Medicine at Mount Sinai Program for the Protection of Human Subjects (PPHS) under study number 20-00341. Data for the analysis including the clinical covariates are available in Synapse syn35874390.^14^ Access to the data and steps to process the clinical information to create the cohort are detailed on the site.^14^ All clinical experimentation methods pursued in this study are in adherence with the Declaration of Helsinki.

**Figure 1a.**
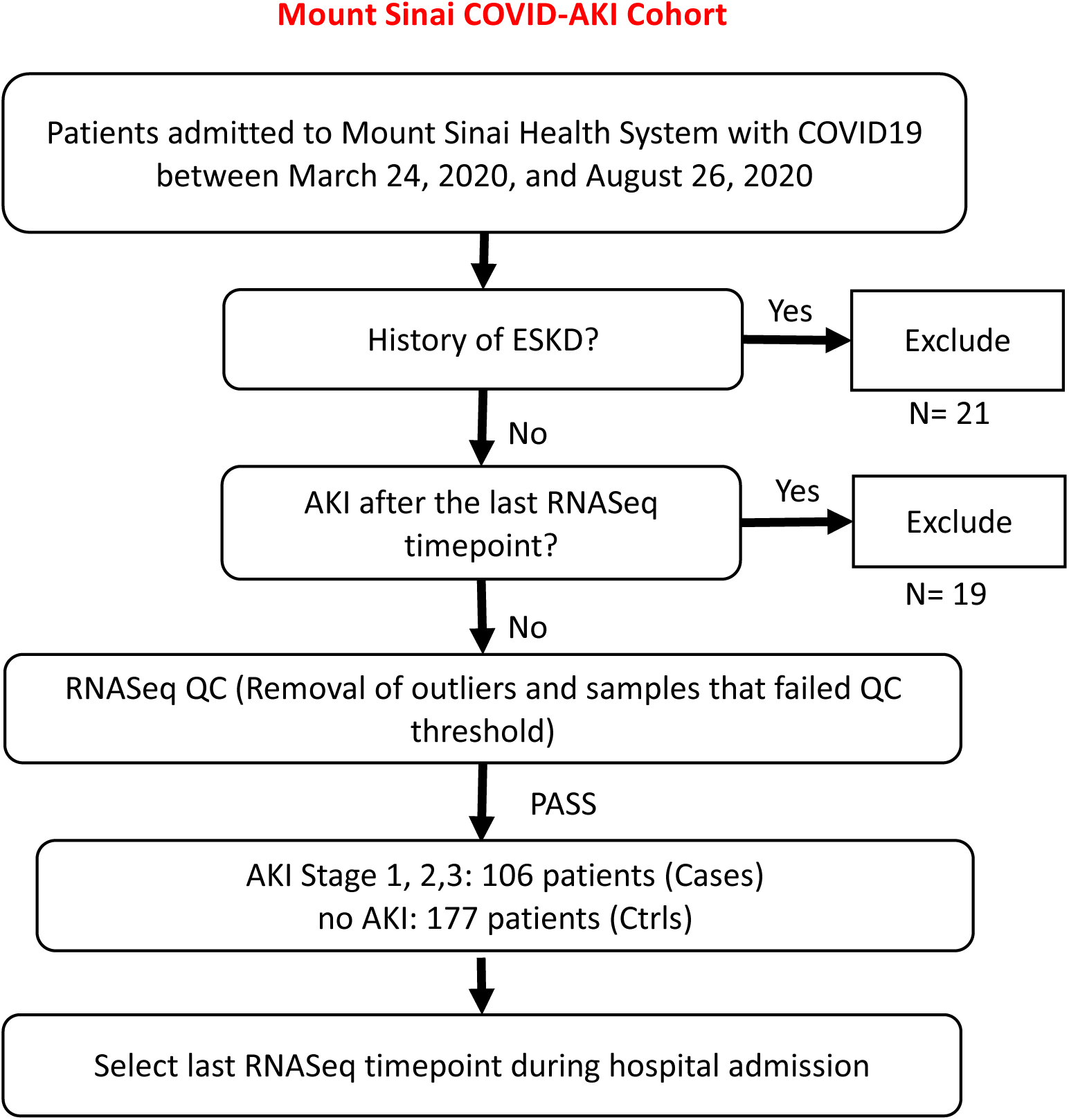
Flowchart of patients included in the current study.

**Figure 1b.**
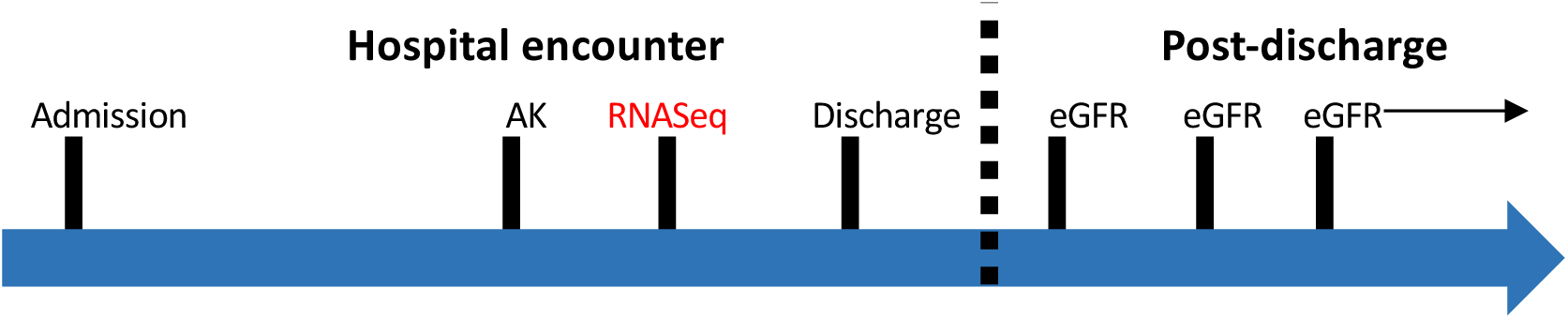
Timeline of sample collection for current study

**Figure 1 Legend.** Figure 1a shows the patients and samples included in the current study, and Figure 1b shows a visualization of the sample collection timeline.

### Definition of Acute Kidney Injury

We defined AKI per Kidney Disease Improving Global Outcomes (KDIGO)^20^ criteria: an increase in serum creatinine by an absolute value of 0.3 mg/dL in a period of 48h or by at least 1.5 times the baseline creatinine (historical measurement) within a period of 7 days. For patients with previous serum creatinine measurements available in the 365 days before admission, we considered the minimum value as the baseline creatinine. For patients without baseline creatinine in this period, a baseline reference value was used based on an estimated glomerular filtration rate (eGFR) of 75 ml/min per 1.73m^2^ was implemented from the MDRD equation, as per the KDIGO AKI guidelines.^21,22^

### Sample Collection, Sequencing, and RNA-seq count data processing

We extracted patient demographic and laboratory data from an institutional electronic health records (EHR) database^14,19^. Further processing of the whole blood and PBMC samples, RNA sequencing, and quantification were carried out according to previously standardized protocols (**Supplementary Information**).^14^ We normalized the raw counts for the gene expressions using the edgeR package and log_2_ transformed to log counts per million (CPM). We then normalized raw count data (58920) using the calcNormFactors function and then transformed to normalized log_2_ CPM with weights computed by *voom* from the *limma* (R v4.3) package^23^. We included genes with more than 1 count per million (CPM) in at least 10% of the samples (28 samples) in the analysis (n=18553).^23^

### Dimensionality Reduction and Linear Model Development

We performed principal component analysis (PCA) (**Supplementary Figure 1**) using *voom*-transformed normalized log_2_ CPM of the genes in the input matrix using prComp function (R v4.0.3) to plot the top 4 principal components (PCs). We then computed canonical correlation among the technical, biological and clinical covariates using CanCorrPairs^24^ from the variancePartition R package and visualized the associations using the plotCorrMatrix function (**Supplementary Information**). The workflow to compute canonical correlations and visualize them was applied in a previously published workflow on samples that were part of the same biobank for a different study (**Supplementary Information**).^14^ Following this step, we undertook a data-driven approach to identify a series of non-redundant technical and biological covariates that accounted for a substantial fraction of unwanted variation in gene expression. We iteratively identified a covariate (biological, clinical, or technical) whose effect was the strongest observed driver of unwanted variance in the cohort. The identified variable, thus being tagged as a driver of unwanted variance, was then added to the linear model. We repeated this procedure iteratively until no further confounding variables were observed to be strong drivers of variance in the data, as shown in **Supplementary Figure 1B**. We adjusted for the selected variables in all subsequent linear modeling. The contribution of each of the covariates to the overall variance in the data was visualized by conducting a variance partition analysis using *variancePartition* R package^24^.

### Differential expression and analysis

We carried out differential expression analysis between cases and controls using *limma* (R v4.3) and adjusted the model for the following clinical covariates: presence or absence of mechanical ventilation, sex, age, and chronic kidney disease (CKD) (as documented comorbidity in the EHR). We used cell-type deconvolution using CIBERSORTX^23,24^ to estimate cell type proportions in each sample using LM22 PBMCs as reference following a previously published workflow^14^ and then iteratively adjusted the linear model for neutrophils, plasma cells, macrophages, and CD4+ memory activated T-cells^25^ (**Supplementary Information)**. After multiple testing correction on the p-values of the genes (Bonferroni-adjusted *FDR*□<□0.05), we plotted the statistically significant genes in a volcano plot to depict the separation between the expression of genes with increased and decreased transcription abundances through log fold change (logFC).

### Comparison with Sepsis associated AKI

To account for differences between sepsis-associated and COVID-AKI, we compared the statistically significant (FDR <0.05) list of differentially expressed genes and canonical pathways enriched for these genes to a previously published dataset (including differential gene expression results, pathways and normalized gene expression) from participants with sepsis and AKI before the COVID-19 pandemic.^25^ We downloaded expression data from GEO for project GSE67401^25^ and obtained the associated clinical phenotypes. We performed differential expression analysis using the DESeq2^26^ package following the protocol as described in the original manuscript^25^ and identified differentially expressed genes (FDR <0.05) between the cases and controls in the sepsis-AKI cohort. After Bonferroni adjustment, we performed a one-sided Fisher’s exact test to identify enrichment and analyze overlap between the genes. We also extracted the published pathways from the manuscript^25^ and performed an overlap analysis to identify common pathways enriched in both datasets.

### Characterization of long-term kidney function using markers of AKI

We used post-discharge creatinine values measured for returning patients to compute estimated glomerular filtration rate (eGFR) values using the CKD-EPI equation.^27^ Clinical data pertinent to returning patients within this cohort including protein measurements were extracted from the EHR starting from the day after discharge with follow-up care through 12/2/2021. Outpatient eGFR measurements from repeated serum creatinine measurements from patients contained at least one outpatient post-discharge eGFR measurement. We included 115 patients who returned for post-discharge clinical care in this cohort. To determine association of overall expression of markers for AKI with long-term post-discharge kidney function, we modeled the association of the differential gene signatures of AKI in COVID-19 to the overall changes in post-discharge eGFR using a mixed-effects linear regression model (lme4^28^ package in R v4.0.3). We adjusted the model for baseline creatinine, number of days since RNA-seq sample was extracted, presence of AKI during hospitalization, and the individual gene expressions as the fixed effect and patient ID as random effect accounting for the correlation among eGFR values from the same individuals. We evaluated significance of the β coefficient for the expression of each of the independent gene expressions using a *t-test* with Satterthwaite degrees of freedom implemented in the *lmerTest* R package.^28^ We adjusted p-values using the Benjamini-Hochberg procedure (FDR < 0.05).^29^ We then plotted the post-discharge eGFR values over time for individuals clustered by gene expression tertiles (bottom 33^rd^ percentile, middle 33^rd^ percentile, and top 33^rd^ percentile) for AKI cases and controls separately (**Supplementary Figure S5**).

### Functional association and statistical analysis

Pathway enrichment analysis was performed for the differentially expressed genes through Ingenuity Pathway Analysis (IPA)^26^ to uncover functional associations and causal networks enriched (Fishers exact test p-value < 0.05) for differentially expressed genes in the COVID-associated AKI cohort. IPA canonical pathway analysis also included calculated z-scores that represented activation (positive z-score) or inhibition state (negative z-score). We selected for the top canonical pathways at a p-value cut-off of 0.05. Further, top regulatory genes, disease functions, and curated toxicity functions were investigated within IPA for both, up, and downregulated genes.

## RESULTS

### Description of the study population

Of 283 patients in the COVID-AKI cohort, 106 (37%) had AKI. Patients with COVID-associated AKI were on average older than those without (67 years vs 60 years, p=0.014) with a similar distribution of race (36% White and 21% Black versus 32% White and 24% Black patients, p-value not significant). Sequential Organ Failure Assessment (SOFA) scores^30^ were also significantly greater in those with COVID-associated AKI compared with controls (5.7 vs 1.2, p<0.001). Laboratory parameters including blood urea nitrogen levels (52.2 mmol/L vs 16.9 mmol/L, p < 0.001), leukocyte counts (12.8×10^9^/L vs 7.8×10^9^/L, p<0.001), and ferritin (1650 ug/L vs 932 ug/L, p 0.027) were significantly higher in patients with AKI. Patients with COVID-associated AKI had a greater prevalence of atrial fibrillation (20% vs 10%, p 0.018), type 2 diabetes (37% vs 20%, p<0.001), and comorbid CKD (20% vs 5%, p<0.001) and were also more likely to have been on any vasopressors (52% vs 7%, p <0.001) during their hospitalization.

The post-discharge eGFR cohort comprised 115 with follow up data (**Table 1B**). 34 had AKI and were part of the COVID-AKI case cohort. Median age was 62 years, 50% were male and 24.3% white. 11% of the patients had CKD stage 3 or higher and 30% had type 2 diabetes. The median duration of follow-up was 162 days with a median of 4 eGFR measurements (**Table 1C**).

**Table 1A.** Summary statistics for the COVID-AKI cohort.

| Cohort Characteristics | Developed AKI<br>(Stage 1/2/3)<br>during<br>hospitalization | No AKI during<br>hospitalization | p-value |
| --- | --- | --- | --- |
|  | N=106 | N=177 |  |
| **Age, mean (SD)** | 66.5 (15.4) | 61.4 (16.6) | 0.01409 |
| **Male, n (%)** | 63 (59%) | 99 (56%) | 0.62006 |
| **Race, n (%)** |  |  |  |
| White | 38(36%) | 57 (32%) |  |
| Black or African American | 22 (21%) | 43 (24%) |  |
| Other | 46 (42%) | 77 (44%) |  |
| **Ethnicity, n (%)** |  |  |  |
| Hispanic or Latino | 50 (47%) | 62 (35%) | 0.03523 |
| Not Hispanic or Latino | 55 (53%) | 110 (62%) | 0.03523 |
| Unknown / Not Reported | 1 (0%) | 5 (3%) |  |
| **Vitals & Lab Parameters during**<br>**Hospitalization, n (SD)** |  |  |  |
| SOFA | 5.79 (4.35) | 1.2 (1.65) | <0.001 |
| Modified SOFA | 4.56 (4.03) | 1.09 (1.52) | <0.001 |
| Baseline Creatinine (mg/dL) | 1.07 (0.592) | 0.929 (0.27) | 0.34942 |
| Maximum Lactate (mmol/L) | 1.88 (1.32) | 1.3 (0.43) | 0.35154 |
| Minimum Systolic Blood Pressure (mmHg) | 106 (17.2) | 111 (13) | 0.00409 |
| Maximum Systolic Blood Pressure (mmHg) | 142 (20.9) | 137 (20.5) | 0.02945 |
| Maximum Pulse (bpm) | 103 (18.5) | 90.5 (17.9) | <0.001 |
| Maximum Blood Urea Nitrogen (mmol/L) | 52.2 (34.6) | 16.9 (10.4) | <0.001 |
| Maximum White Blood Cell Count (10<sup>9</sup>/L) | 12.8 (7.19) | 7.8 (3.46) | <0.001 |
| Minimum Platelet Count (10<sup>9</sup>/L) | 270 (144) | 318 (140) | 0.00664 |
| Minimum Lymphocyte count (10<sup>9</sup>/L) | 2.03 (3.17) | 2.31 (3.71) | 0.00862 |
| Maximum Creatinine (mg/dL) | 2.71 (2.6) | 0.862 (0.365) | <0.001 |
| Maximum Ferritin (ug/L) | 1650 (2140) | 932 (1240) | 0.02712 |
| Maximum IL1-β (pg/mL) | 0.4 (NA) | 0.6 (0.216) | 0.46816 |
| **Comorbidities, n (%)** |  |  |  |
| Atrial Fibrillation | 21 (20%) | 17 (10%) | 0.01893 |
| Coronary Artery Disease | 19 (18%) | 26 (15%) | 0.50390 |
| Arterial Hypertension | 48 (45%) | 71 (40%) | 0.45556 |
| Diabetes | 39 (37%) | 32 (18%) | 0.00063 |
| Chronic Kidney Disease | 21 (20%) | 8 (5%) | <0.001 |

Peripheral Transcriptomics in Acute and Long-Term Kidney Dysfunction in SARS-CoV2 Infection
| **Highest Respiratory support n(%)** |  |  |  |
| --- | --- | --- | --- |
| Intubation | 49 (46%) | 9 (5%) | <0.001 |
| Non-Invasive Ventilation | 9 (8%) | 8 (5%) | 0.20040 |
| Nasal Cannula | 34 (32%) | 91 (51%) | 0.00194 |
| No Ventilation | 14 (13%) | 69 (39%) | <0.001 |
| **Vasopressor Use during Hospitalization n (%)** |  |  |  |
| Any vasopressor | 55 (52%) | 12 (7%) | <0.001 |
| Norepinephrine | 49 (46%) | 10 (6%) | <0.001 |
| Vasopressin | 18 (17%) | 2 (1%) | <0.001 |
| Phenylephrine | 15 (14%) | 1 (1%) | <0.001 |
| Epinephrine | 3 (3%) | 2 (1%) | 0.36667 |
| Milrinone | 3 (3%) | 1 (1%) | 0.14950 |
| Dopamine | 1 (1%) | 0 (0%) | 0.37456 |

**Table 1B.** Patient cohort summary statistics for the post-discharge follow-up cohort for long-term eGFR analysis.

| Cohort Characteristics | Developed AKI (Stage 1/2/3) during hospitalization | No AKI during hospitalization | p-value | Total (Category %) |
| --- | --- | --- | --- | --- |
|  | N=31 | N=84 |  | 115 |
| **Age, mean (SD)** | 60 (15.9) | 59 (15.76) |  |  |
| **Male, n (%)** | 16 (52%) | 44 (52.3%) |  | 60 (52.17%) |
| **Race, n (%)** |  |  |  |  |
| White | 7(22.5%) | 21 (25%) |  | 28 (24.34%) |
| Black or African American | 7 (22.5%) | 25 (29.76%) |  | 32 (27.82%) |
| Other | 17 (54.83%) | 38 (45.23%) |  | 55 (47.82%) |
| **Ethnicity, n (%)** |  |  |  |  |
| Hispanic or Latino | 14 (45.16%) | 27 (32.14%) |  | 41 (35.65%) |
| Not Hispanic or Latino | 16 (51.61%) | 50 (59.52%) |  | 66 (57.39%) |
| Unknown / Not Reported | 1 (0%) | 7 (1%) |  | 8 (7%) |
| **Vitals & Lab Parameters During Hospitalization, n** |  |  |  |  |

Peripheral Transcriptomics in Acute and Long-Term Kidney Dysfunction in SARS-CoV2 Infection
| (SD) |  |  |  |  |
| --- | --- | --- | --- | --- |
| SOFA | 3.84 (2.72) | 1.17 (1.82) | p < 0.001 |  |
| Modified SOFA | 2.84 (2.78) | 1.05 (1.66) | p < 0.001 |  |
| Baseline Creatinine (mg/dL) | 1.17 (0.6) | 0.968 (0.211) |  |  |
| Maximum Lactate (mmol/L) | 1.65 (1.19) | 1.7 (0.283) |  |  |
| Minimum Systolic Blood Pressure (mmHg) | 109 (16.4) | 112 (12.9) |  |  |
| Maximum Systolic Blood Pressure (mmHg) | 137 (17.2) | 138 (20.8) |  |  |
| Maximum Pulse (bpm) | 98.8 (16.3) | 89.5 (14.2) | p = 0.0023 |  |
| Maximum Blood Urea Nitrogen (mmol/L) | 41.9 (28.2) | 16.3 (9.41) | p < 0.001 |  |
| Maximum White Blood Cell count (10<sup>9</sup>/L) | 9.46 (5.16) | 7.62 (3.25) |  |  |
| Minimum Platelet count (10<sup>9</sup>/L) | 252 (150) | 328 (151) | p = 0.00823 |  |
| Minimum Lymphocyte count (10<sup>9</sup>/L) | 2.97 (4.06) | 2.59 (4.45) |  |  |
| Maximum Creatinine (mg/dL) | 2.27 (2.11) | 0.894 (0.408) | p < 0.001 |  |
| Maximum Ferritin (ug/L) | 1900 (2810) | 981 (1380) |  |  |
| Maximum IL-1β (pg/mL) | 0.4 (NA) | 0.7 (0.283) | | |
| **Comorbidities, n (%)** |  |  |  |  |
| Atrial Fibrillation | 8 (26%) | 11 (13%) |  | 19 (16.52%) |
| Coronary Artery Disease | 8 (26%) | 13 (15%) |  | 21 (18.26%) |
| Arterial Hypertension | 15 (48%) | 40 (48%) |  | 55 (47.82%) |
| Diabetes | 14 (45%) | 21 (25%) | p = 0.043 | 35 (30.43%) |
| Chronic Kidney Disease | 8 (26%) | 5 (6%) | p = 0.00587 | 13 (11.3%) |
| **Vasopressor use during hospitalization. n (%)** |  |  |  |  |
| Any Vasopressor | 15 (48%) | 7 (8%) | p < 0.001 | 22 (19.13%) |
| Norepinephrine | 12 (39%) | 6 (7%) | p < 0.001 | 28 (24.34%) |
| Vasopressin | 7 (23%) | 1 (1%) | p < 0.001 | 8 (7%) |
| Phenylephrine | 2 (6%) | 1 (1%) |  | 3 (3%) |
| Epinephrine | 0 (0%) | 2 (2%) |  | 2 (1.7%) |
| Milrinone | 2 (6%) | 1 (1%) |  | 3 (3%) |

**Table 1C.**
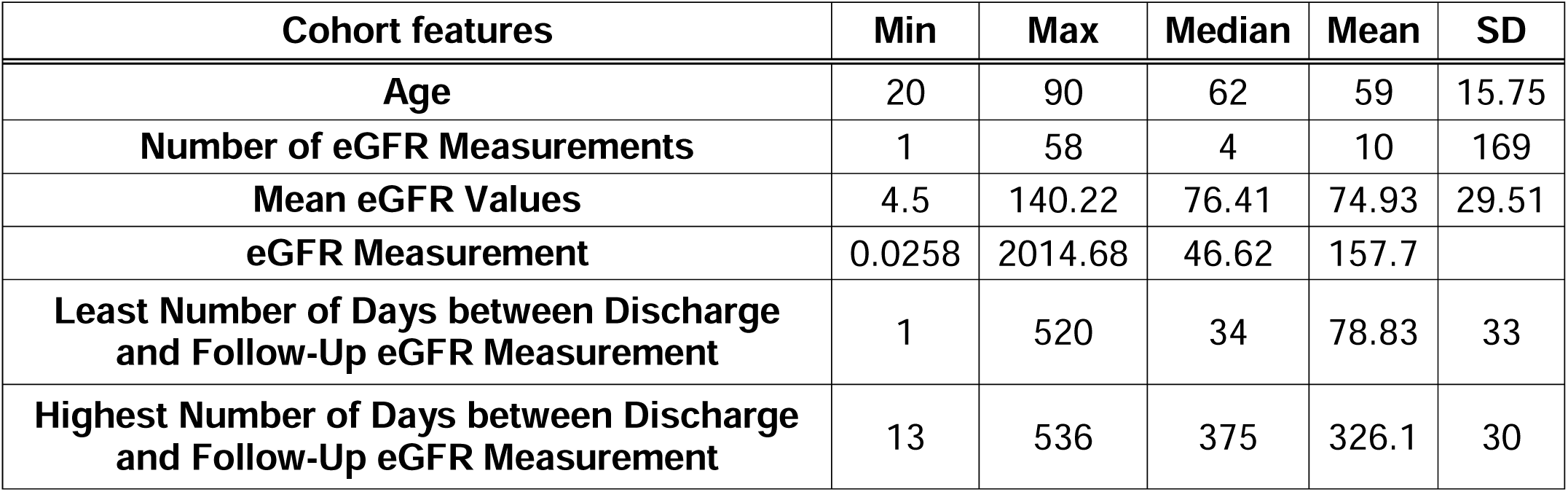
Additional summary statistics for the long-term eGFR patient cohort in Table 1B.

### Differential Expression Analysis

Differential expression analysis yielded 2635 genes which were differentially expressed in cases: 1223 higher expression and 1412 genes with lower expression in AKI versus control (FDR<0.05) (**Figure 2A, Supplementary Table 2A & 2B**).

**Figure 2a.**
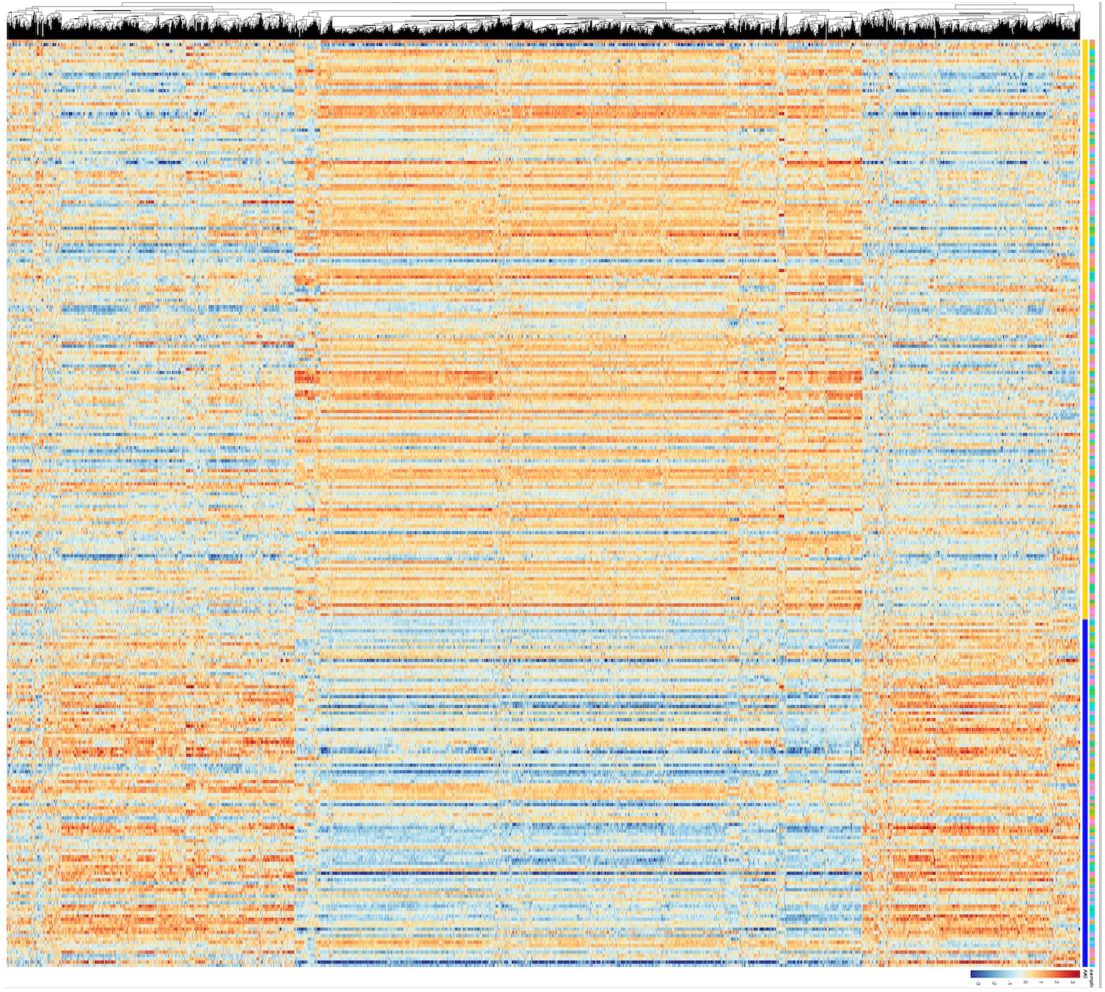
Heatmap showing the differential expression of 2635 genes between cases (COVID-AKI) and controls (COVID with no AKI).

**Figure 2b.**
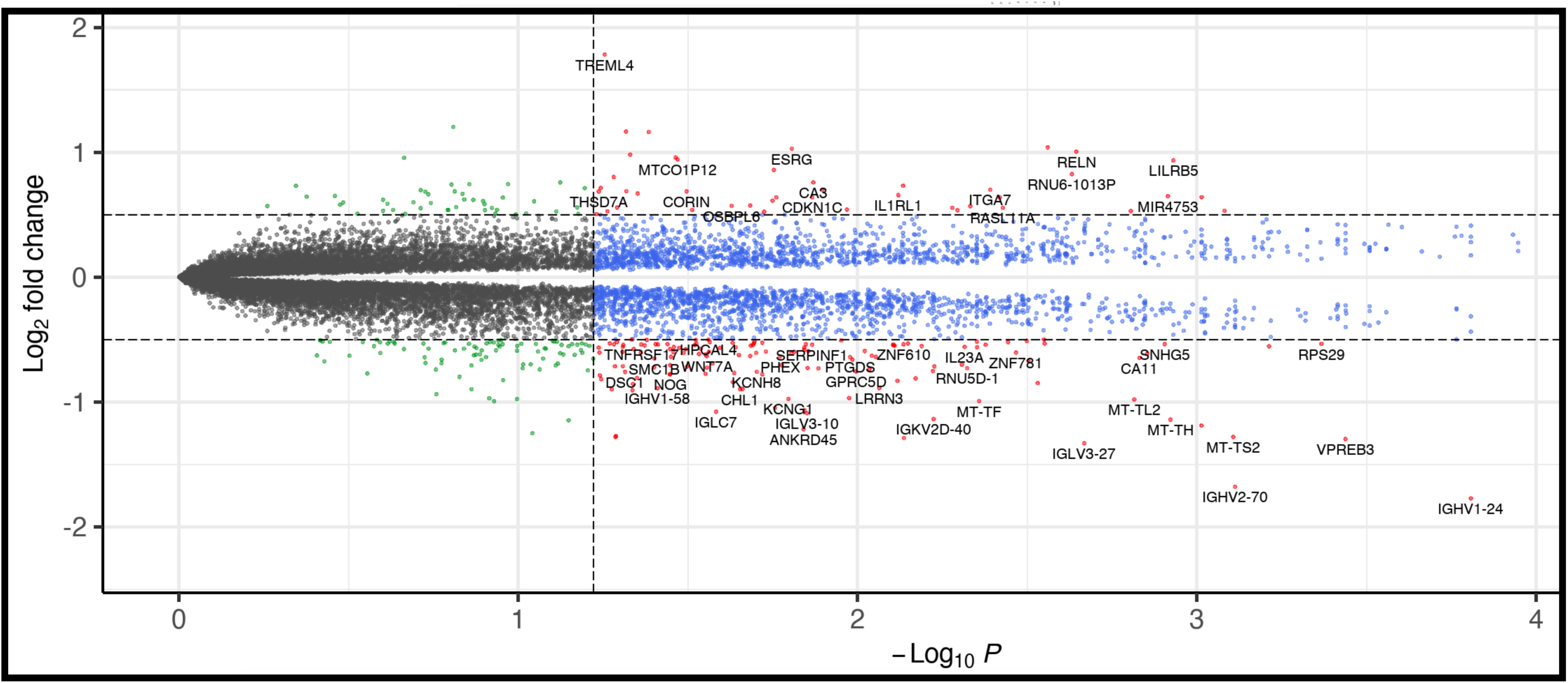
Volcano plot of the differentially expressed genes showing large numbers of significantly downregulated mitochondrial toxicity and endoplasmic reticulum genes.

Functional analysis of the differentially expressed genes revealed a subset of genes involved in upstream regulation of transcription targets (regulatory genes) were also significantly differentially expressed. Master regulators, namely, *mir-21, KLF6, MEG3, KIT, PRL and TGFβ1* were overexpressed (∼1.5x fold increase), while *IFN-gamma (IFNγ or IFN-g), mir-15, mir-19, CAB39L* and *EIF3E* had significantly lower levels of gene expression than controls (∼0.5x fold decrease). Of the known biomarkers for AKI progression and renal tubular injury *HIF1A*, *KLF6, IL1-R1* and *mir-21* showed significantly increased expression (**Supplementary Table 2C**).

### Functional analysis of differentially expressed genes

Functional enrichment analysis identified several enriched pathways associated via an overlap with the expressed genes in the dataset (right-tailed Fisher’s exact test P-value < 0.05, Benjamini Hochberg correction) (**Figure 3 and Supplementary Information Figure S2A-C**). ‘Canonical pathways’ are identified as well-characterized and hand-curated signaling and metabolic pathways that have been extracted from literature, textbooks, and the HumanCyc database, maintained in IPA. The top pathways were *EIF2* signaling/regulation of *eif4* and *p70S6K* signaling, coronavirus pathogenesis pathway, oxidative phosphorylation, mitochondrial dysfunction, and endoplasmic reticulum (ER) stress (**Figure 3A**). Pathway overrepresentation analysis also interrogated the differentially expressed genes in IPA-curated pathways (**Supplementary Information Figure S2B**). Among them, ‘mitochondrial dysfunction’ and ‘*NF-κB* signaling’ had large percentages of downregulated genes, while pathways that impacted cardiac hypertrophy, increased renal damage, and *PPAR/RXR* activation had higher percentages of upregulated genes.

**Figure 3a.** Canonical pathways identified in IPA during the functional characterization of the differentially expressed genes. The top pathways resulting from the analysis include *EiF2* signaling, *eIF4/p70S6K* signaling, oxidative phosphorylation and mitochondrial dysfunction.

**Figure. 3b.** Curated list from Ingenuity Pathway Analysis

**Figure 3 Legend.**
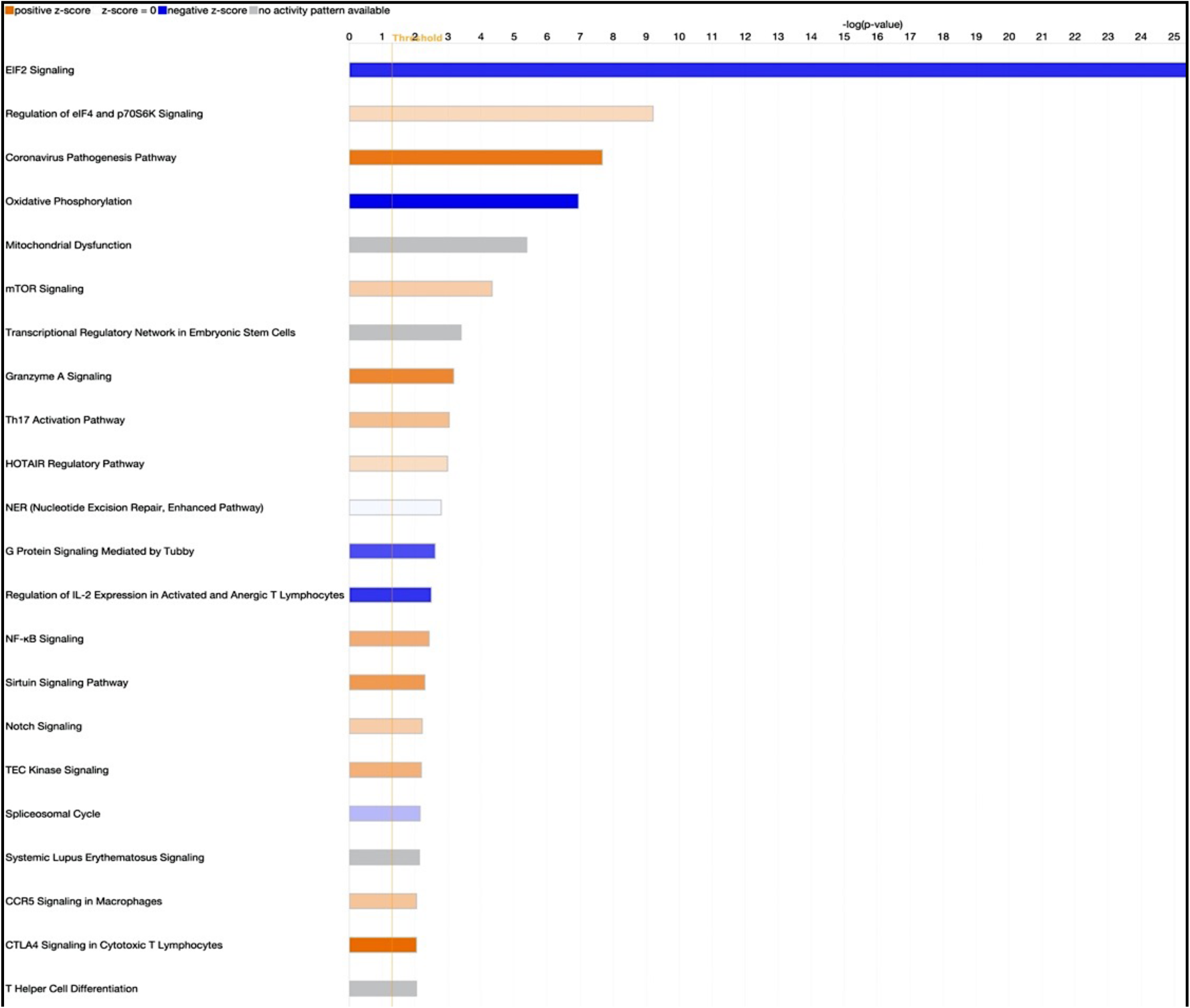
Figure 3b shows a majority of downregulated genes in ‘mitochondrial dysfunction’ and ‘*NF-*K*B* signaling’. Additionally large number of upregulated genes are involved in the pathways involved in ‘cardiac hypertrophy’, ‘increased renal dysfunction’ (PPARα activation) and damage.

### Comparison with sepsis associated AKI

Of the published canonical pathways in the s-AKI dataset, more than 50% of the pathways overlapped with the enriched canonical pathways in the COVID-AKI dataset **(Supplementary Table 2.C.)**. Cellular processes associated with mitochondrial dysfunction, oxidative phosphorylation, and cellular stress (see **Supplementary Fig S4**) were some of the top common pathways. Of the 630 significant differentially expressed genes generated from the s-AKI cohort, 111 of them were also found in the COVID-AKI dataset. The Fisher exact test statistical significance for enrichment of sepsis-AKI genes in COVID-AKI cohort was < 0.00001 (**Supplementary Information**).

### Characterization of post-acute kidney dysfunction

COVID-19 is associated with long-term eGFR decline.^13,15^ Of the ∼2619 differentially expressed genes for COVID-AKI, we also found 164 genes **(Figure 4)** were associated with overall change in eGFR (**Supplementary Table 2D & 2E**). Those with the highest ranked association (ranks determined by the strength of the association **(Figure 5)** of the gene expressions against overall rate of eGFR) are *CALCOCO2, SIK3, mir29, KDM8,* and *MEF2C* (**Supplementary Table 2F**). Pathway over-representation analysis in IPA revealed an enrichment for ABRA signaling pathway with MEF2C and TAGLN as key regulators in the pathway (**Supplementary Table 2G**). Significant functional and diseases associations in IPA included regulation of actin cytoskeleton, cellular adhesion and proliferation, and cardiovascular system development and function (IPA Content Version: 101138820, Release Date: 2023-08-24).

**Figure 4.**
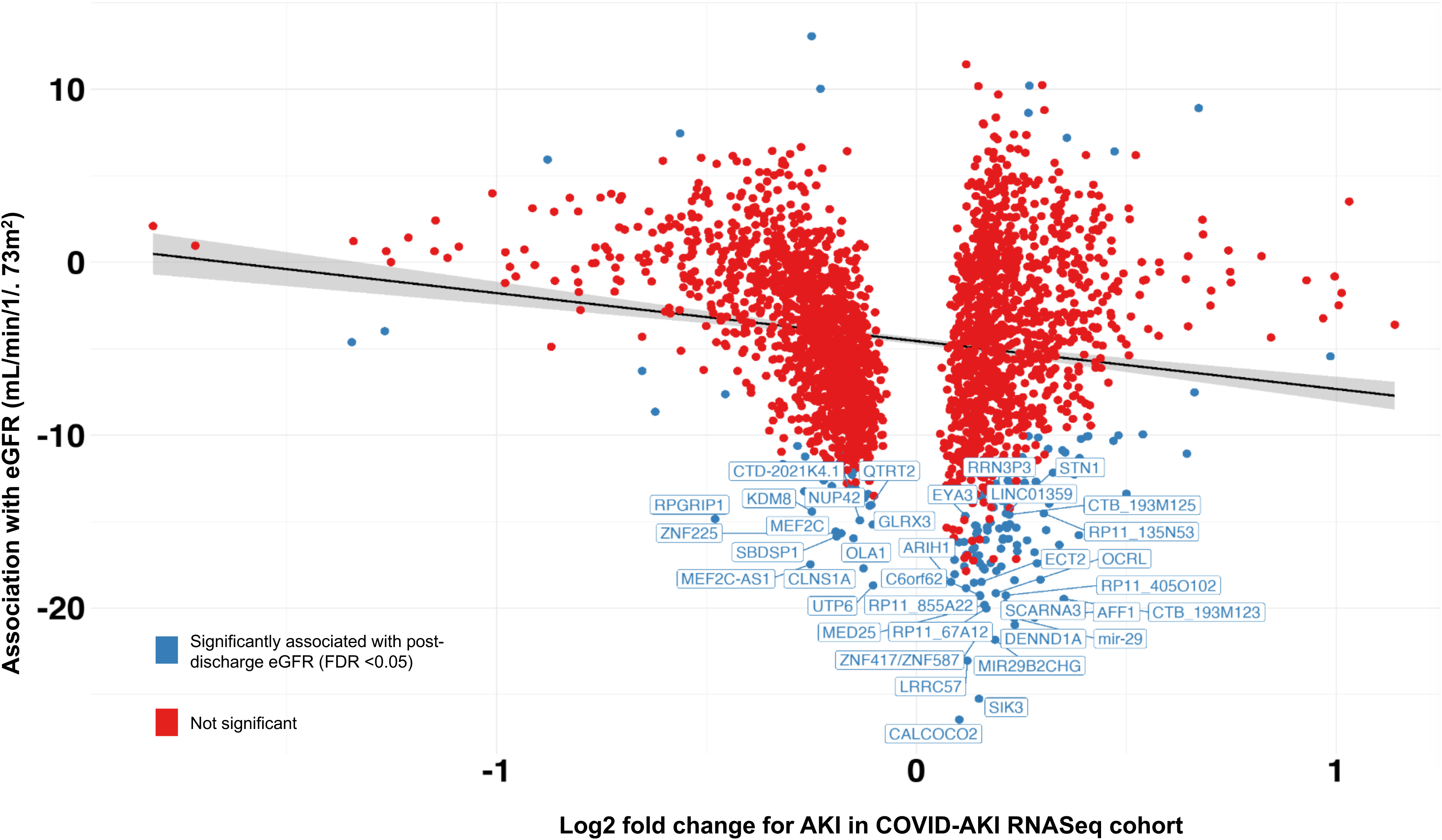
Association of differentially expressed gene signatures in AKI in the COVID-AKI cohort and overall decline in eGFR in the long-term cohort 1-year post-discharge.

**Figure 4 Legend.** Figure 4 shows a plot of the expression of 2635 genes associated with AKI in the COVID-AKI cohort against the β estimates of the linear mixed model that analyzes the effect of these expressions over the long-term decline of eGFR. The gene expressions are plotted by fold change on the x-axis (log2 fold change). The y-axis is the beta estimate of the overall change in long-term eGFR from the linear mixed model analysis. A negative beta(β) estimate for overall change in long-term eGFR, signals that genes that show a higher positive expression are negatively correlated with overall decrease in post-discharge eGFR while genes with a higher negative expression are directly correlated with overall decrease in post-discharge eGFR.

## DISCUSSION

To our knowledge, this is the largest peripheral transcriptomics study in hospitalized COVID-19 patients who develop AKI. Our results show that COVID-AKI is associated with molecular perturbations within the regulatory markers of endoplasmic reticulum (ER) stress,^31^ tubular injury, mitochondrial dysfunction, and oxidative phosphorylation. Some pathways are also found to be common to those with sepsis-AKI including Hypoxia Signaling in the Cardiovascular System, ERK/MAPK Signaling, Unfolded protein response, and *IL-1* signaling.^32–34^ In addition, we uncover associations between a subset of these acute-phase, COVID-19 molecular markers and long-term kidney function dysfunction, including markers of autophagy, renal impairment via fibrosis, and cardiac structure/function.

We utilized peripheral blood mononuclear cells (PBMCs) as a model system to understand the underlying peripheral molecular mechanisms of AKI in COVID-19. One example of the mononuclear cell system is circulating monocytes, influenced by the cytokines or other molecular interactions to be recruited into injured tissues to differentiate into specific macrophage phenotypes.^35^ Thus, PBMCs serve as surrogates for systemic physiological changes,^36,37^ and their bioenergetic profiles have gained substantial attention^38^ in recent investigations of various diseases.^39^

The *eiF2* pathway was inhibited, *and mTOR* was activated in our cohort (**Figure 3A**). These pathways have previously been implicated in AKI development both in ischemia-reperfusion and in sepsis-associated AKI. Coronaviruses induce cellular stress by disrupting cellular homeostasis and triggering ER stress^40,41^ and cellular stress induced activation of *eiF2-α* kinases and phosphorylation of the α subunit of *eIF2* to inhibit *eIF-2* signaling.^42^ Additionally, viral infection acts via the *eIF2α* pathway to cause global inhibition of translation while upregulating proinflammatory cytokines.^31,42^ The *mTOR* pathway is intertwined with the activation of ER stress response via eukaryotic initiation factor (*eIF2/4* complex).^43,44^ As a viable target for therapeutic intervention, rapamycin, an inhibitor of mTOR signaling, has been shown to offer a protective effect against progression in rodent models of CKD.^45^ Other *mTOR* inhibitors (everolimus and sirolimus) used in immunosuppression in transplant patients have alternatively been shown to contribute to AKI progression and tubular injury.^46^ This suggests that *mTOR* may initially play a role in AKI pathogenesis but inhibition of the appropriate targets within the signaling pathway might prevent the progression of AKI to CKD. We also find that most mitochondrial genes are downregulated in COVID-19 patients with AKI. Mitochondrial oxidative phosphorylation pathways play a key role in the production of ATP and adaptive response to systemic inflammation, oxidative stress, and prevention of tubular injury.^47^ Inhibition or downregulation of these pathways would likely contribute to maladaptive stress response and extended ischemia. Therapies targeting mitochondrial dysfunction, including pharmacological (SS-31,^48^ mitoQ^49^), cellular, and even mitochondrial transplantation, have potentially been considered for the treatment of ischemia-reperfusion injury. Given that mitochondrial dysfunction and downregulation of oxidative phosphorylation are key in mediating COVID-associated AKI^50^ and CKD,^51^ one should consider repurposing therapies targeting mitochondria for the mitigation of kidney failure. In addition to inflammation and cellular stress responses/renal tubular injury, some of the molecular biomarkers have also previously been implicated in the worsening of renal damage in COVID-AKI patients. Specifically, genes such as *mir-21 and HIF-1 alpha* (*HIF-1a*) have been known to play a significant role in the pathogenesis and progression of AKI to chronic renal disease. MicroRNAs are known to be critical in the activation of the innate immune system and the regulation of the adaptive immune response.^52^ Mir-21 is a known regulator of apoptosis and involved in inflammatory and signaling pathways that may lead to hypoxia and tubular damage in patients with AKI.^52^ In another study, in PBMCs, mir-21 were shown to contribute to the underlying atherosclerosis by participating in the inflammatory processes governing angiogenesis in coronary artery disease.^53^ HIF-1 alpha (HIF-1a or HIF1a) is also known as a master regulator of adaptive immune responses to hypoxia, a common condition in macrophage-driven inflammation due to a condition such as COVID-19.^54^ In addition, HIF-1a and its alternate isoform, HIF-2a exert mutually antagonistic effects on the hypoxic states of the cell and the inflammatory pathways, which causes a reduction of Sirt-1 thus leading to chronic heart failure in the long term.^55^ Furthermore, *EIF2* and *mTOR* pathways are also known to play a role in both autophagy and fibrosis, making them ideal therapeutic targets for the prevention of COVID-associated AKI progression or PASC.^42,56^ These instances of cardiorenal crosstalk play a common role in COVID-19 AKI and drive the progression to CKD in the long-term.^57^

Previous studies have shown through histopathology and gene expression studies that there are significant similarities in the morphological and molecular profiles of patients with COVID-19-AKI and sepsis-associated AKI.^58,12^ In comparing our data from a pre-COVID-19 sepsis-associated AKI cohort,^25^ we found that of the 55 reported pathways, 50% were common to both COVID-associated AKI and sepsis-associated AKI. The chief pathways that overlapped (**Supplementary Table 2C** and **Supplementary Figure 4A**) included pathways involved in inflammation, kidney injury, and mitochondrial response, namely ‘T_reg_ signaling’, ‘ERK/MAPK signaling’, ‘IL-1 signaling’, ‘hypoxia signaling in the cardiovascular system’, ‘unfolded protein response’, and ‘RAR activation’. Pathways related to renal inflammation, tubular injury, and oxidative phosphorylation were implicated in the pathogenesis of AKI and progression to CKD in both this transcriptomic analysis and in our previously published proteomic analyses.^17^ Our results indicate that therapies that target the immune system in sepsis-associated AKI could be of potential therapeutic advantage in COVID-AKI.

COVID-19 is associated with long-term kidney dysfunction, especially after AKI^13,16^. Our analysis showed that 164 genes (**Table 2D-E**) from the 2635 DE genes implicated in the acute phase are associated with the overall trend of post-discharge progressive loss of renal function (**Table 2F**). We highlight the top 6 genes (ranked by strength of association and fold change) (**Figure 4**) as potential markers that may enable early risk stratification. Gene expression levels of *CALCOCO2, SIK3*, and *mir-29* were inversely correlated with overall trend of long-term eGFR, while gene expression levels of *KDM8*, and *MEF2C* were positively correlated with the overall trend of long-term eGFR. Calcium binding and coiled-coil domain-containing protein 2 (*CALCOCO2*, commonly known as *NDP52)* is an autophagy-related gene. In PBMCs interferon-signaling factor *tetherin* regulates mitophagy^59^ via activation of NDP52. In patients with dilated cardiomyopathy, *NDP52* expression associated with autophagy-related reduced left ventricular function^60^. *SIK3,* a regulator of pro-inflammatory cytokines such as TNF-α,^61^ also polarizes macrophages leading to fibrosis and scarring^35,61^ in smooth muscle tissues.^62^ Mir-29 is a master regulator of the adaptive immune system and contributes to the pathophysiology of CKD^63,64^ via a reno-protective effect.^65^ In addition, *mir-29* plays a role in cardiac remodeling and hypertrophy^66^ and is viewed as a potential biomarker for early detection and progression of CKD.^64^ Loss of *MEF2C* in B-cells^67^ causes defects in b-cell proliferation and survival via p38/MAPK pathway^68^ and is prognostic of CKD progression in elderly patients.^69^ *KDM8* (*JMJD5),* a critical regulator of DNA replication^70^ under DNA damage stress,^71^ is shown to epigenetically modulate cardiac metabolism via the p53/NFKB activation.^72^ In patients with diabetes, downregulation of *KDM8* was associated with the risk of dilated cardiomyopathy (DCM).^73^ In addition, pathway overrepresentation analysis also highlighted the Actin-binding Rho activating (ABRA) signaling pathway (**Supplementary Table 2G**) in and the association of Rho GTPases with cardiac structure and function^74^ and regulation of renal physiology and nephropathies.^75^

A unique aspect of our study is the diversity of our patient cohort. The patient cohort for the COVID-19 AKI study also reflects the distribution of race and ethnicity^76^ of the NYC catchment area.^77^ The overall racial and ethnic distribution improves the generalizability and application of our findings to diverse populations.

It is important to interpret our results in the context of certain limitations. First, samples were collected during the hospital course of patients with confirmed COVID-19. However, logistical challenges during the pandemic resulted in the samples that were not collected at the exact same number of days post-hospitalization and were not standardized between patients. In addition, the time since the start of the SARS-CoV2 infection is unknown. This scenario is true across all COVID datasets and is not unique to ours. It is also important to note that while the samples were collected at multiple time points, our analysis was conducted using gene expression measurements from a single time point, after the patients had their last instance of AKI during hospitalization. Second, all patients enrolled in our study were collected in 2020, while our sepsis-associated AKI cohort was derived from a study in 2015 with different sample processing and library preparation methodologies before RNA-seq. While variability in processing strategies may reduce signal-to-noise when comparing the two cohorts, the different time periods of collection ensured that neither cohort has any confounding from possible SARS-CoV2 infection/hospital-acquired SARS-CoV2 infections. Third, our current cohort was enrolled in the early pandemic between March-August of 2020. Given the large sample size of our study complicated by SARS-COV2 strain evolution via dynamic positive selection of mutations within the spike protein rather than mutations in the core,^78,79^ these results likely generalize across different variants. It would be remiss to not mention the subjective nature of interpretation of peripheral blood RNASeq results as a heterogenous population can be affected by changes in the percentage of cell types. Future directions necessitate the need for single-cell RNASeq studies to disentangle these effects effectively and clearly.

In conclusion, we present a whole blood transcriptomic comparison of COVID-19 patients with and without AKI and long-term kidney dysfunction in the largest sample size to date. These results may have clinical utility in risk stratification of long-term kidney dysfunction after COVID-19 and for therapeutic repurposing.

## Data Availability

Data Availability
This research was reviewed and approved by the Icahn School of Medicine at Mount Sinai Program for the Protection of Human Subjects (PPHS) under study number 20 00341. The clinical data tables and analysis data are available in the Synapse repository syn35874390. Synapse can be accessed at https://www.synapse.org/#!Synapse:syn35874390.
Subject IDs in the Supplementary Tables are public deidentified ids from the synapse repository

## ACKNOWLEDGEMENTS.

We thank Wei Guo, Lili Gai, and Eugene Fluder of the High-Performance Computing group at 373 Mount Sinai for enabling the computational infrastructure underlying these experiments.

## AUTHOR CONTRIBUTIONS

Conceptualization: GNN, PJ, IP.

Methodology: PJ, IP, LL, RT, NB, MSF, GVR, SD, SKS, EK.

Investigation: PJ, IP, RT, DMDV, GVR, SD, EGK, FG, JK, AC.

Visualization: PJ, IP, MSF, WO, JK, RT, EGK, AV, AC.

Funding acquisition: GNN, SGC, JCH.

Supervision: GNN, AWC, NB, RT, NB, SGC, LC, JCH.

Writing: PJ, IP, MR, GNN.

Revisions: PJ, MR, ET, RLL, MSF, RT, LC, SD, JJ, FG, GNN.

## DATA SHARING STATEMENT

### Data Availability

This research was reviewed and approved by the Icahn School of Medicine at Mount Sinai Program for the Protection of Human Subjects (PPHS) under study number 20–00341. The clinical data tables and analysis data are available in the Synapse repository syn35874390. Synapse can be accessed at https://www.synapse.org/#!Synapse:syn35874390.

### Code Availability

The code is available at our GitHub repository **Nadkarni-Lab: aki_covid_transcriptomics** here: https://github.com/Nadkarni-Lab/aki_covid_transcriptomics.

