## Supplementary Information for "Peripheral Transcriptomics in Acute and Long-Term Kidney Dysfunction in SARS-CoV2 Infection"

### METHODS

#### Sample Collection, Processing and RNA sequencing

Blood collection was performed along with routine blood draws from consented participants through the length of their hospitalization. Whole blood for RNA-seq was collected in Tempus RNA Blood Tubes (Thermo Fisher Scientific, 4342792). Tubes were then shaken and stored at -80C with minimum delay encountered between blood collection and storage. To account for batch effects, eight samples, believed to be a representative subset of the full cohort for key technical and biological attributes, were manually chosen as “batch controls” and included in every sequencing batch of 192 samples. The detailed sample collection, sample processing and RNA Sequencing protocol has been documented *(Thompson, R.C., et al. Nat Med 2023*) in a prior publication for a larger cohort of which, these samples were a subset. Batch randomization was performed during RNA sequencing and extraction (batch size of 8 within each sequencing batch of 192). RNA extraction, library preparation and sequencing were performed as described in a previous publication *(Beckmann ND, et al. Downregulation of exhausted cytotoxic T cells in gene expression networks of multisystem inflammatory syndrome in children.  Nat Commun 2021)*. We normalized the raw counts for the gene expressions using the edgeR package and $\log_2$ transformed to log counts per million (CPM). We then normalized raw count data (58920) using the calcNormFactors function and then transformed to normalized log2 CPM with weights computed by *voom* from the *limma* (R v4.3) package. We included genes with more than 1 count per million (CPM) in at least 10% of the samples (28 samples) in the analysis (18553).

#### RNA-seq Alignment and Quantification

Post quality-control check, base calls were converted into raw reads and filtered based on read quality. The filtered reads were then aligned to GrCh38 assembly (GENCODE v30 STAR v2.7.3a) using parameters specified in a prior publication (*Thompson, R.C., et al. Nat Med 2023*). Sample mis-labeling correction and sample QC was performed with the removal of samples that did not pass the DV200 (percentage of fragments greater than 200 nucleotides) threshold of 80% or read count below 10 million mapped reads (as mapped by featureCounts from the Subread R package version 1.6.3 and strandness option -s 2 grouped by gene).

#### RNA-seq Count data Processing and Normalization

Counts for all annotated globin genes (gene symbols *CYGB*, *HBA1*, *HBA2*, *HBB*, *HBD*, *HBE1*, *HBG1*, *HBG2*, *HBM*, *HBQ1*, *HBZ* and *MB*) were discarded and the rest of the count matrix was transformed to counts per million (CPM). Genes with CPM ≥ 1 in greater than 10% of the samples in the cohort were included in the analyses. Normalized gene expression values were obtained using trimmed mean of M-values method implemented by the calcNormFactors function in the edgeR package and transformed into normalized $\log_2$ CPM with weights calculated using the voom package.

The final matrix before applying the linear model consisted of ~18500 gene expression CPM for 238 samples.

#### Cell Type Deconvolution and selection for the linear model.

Cell type fractions were estimated for each sample using CIBERSORTx as described in the Methods “Cell type deconvolution and validation” section by Thompson, R.C., et al. Nat Med 2023. Just as previously described, we then used variancePartition and canCorrPairs to determine the correlation of the cell-types to each of the PCs and the outcome. Samples in the cohort that did not pass QC for cell type fractions were removed from the cohort leading to a cohort of 283 samples.

#### Developing the linear model for RNA-Seq analyses.

The existence of multiple technical and biological sources of variation in RNA-seq data often necessitate the exploration of this variance using multiple visualization tools. Principal Component Analysis (PCA) was performed using prComp function in R to identify the separability of the expression of the genes among the cases and controls. Canonical correlations among all technical, clinical, and demographic features were calculated using the canCorPairs function and visualized using the plotCorreMatrix function from the variancePartition R Bioconductor package. An iterative process then ensued starting from the normalized counts to identify the variable that was the most correlated with the top 4 principal components (PCs) relative to its correlation with the outcome. The identified variable, thus being tagged as a driver of unwanted variance was then added to the linear model and this procedure was iteratively repeated until no more confounding variables were observed as strong drivers of variance in the data.

PCA was then performed on the residual expression matrix after applying the linear model and outliers were removed if they were beyond 3 standard deviations outside of the ellipse drawn around the plotted PCs 1 and 2 centered at the origin. Batch effects were adjusted for by including the variable for library prep plate.

#### Differential Expression Analysis

Voom from the Limma package was used to run the differential expression analysis

### RESULTS

#### Principal Component Analysis

**Fig S1.A.**

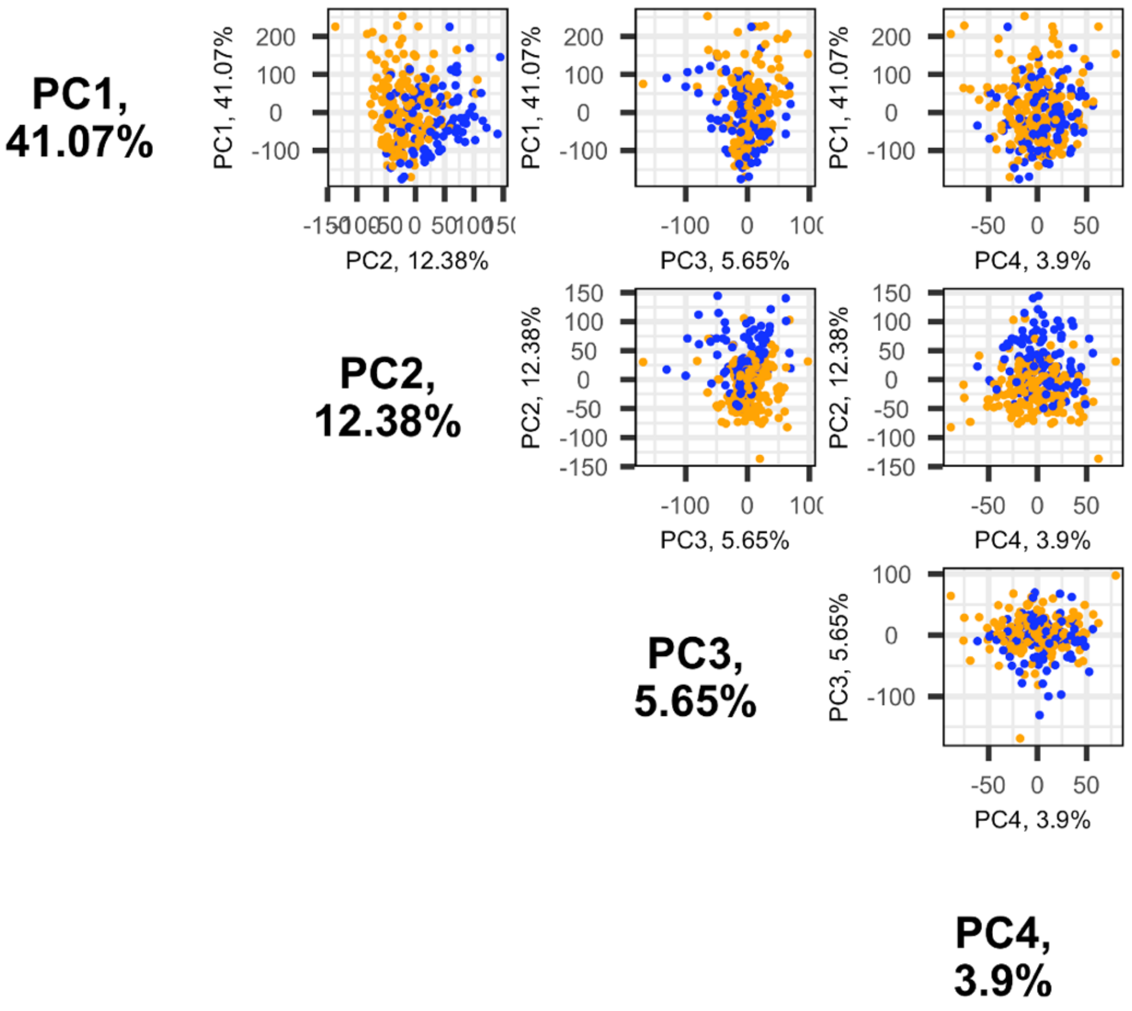

**Fig S1.A.** *Principal Component Analysis* (PCA) of the raw gene expression counts show possible separation along PC2 and PC3. **Fig S1.B.** Visual Representation of the clinical, biological and technical covariates in the linear model and the amount of variation they explained in the data. Plotted using the *variancepartition* function by *Dream* package.

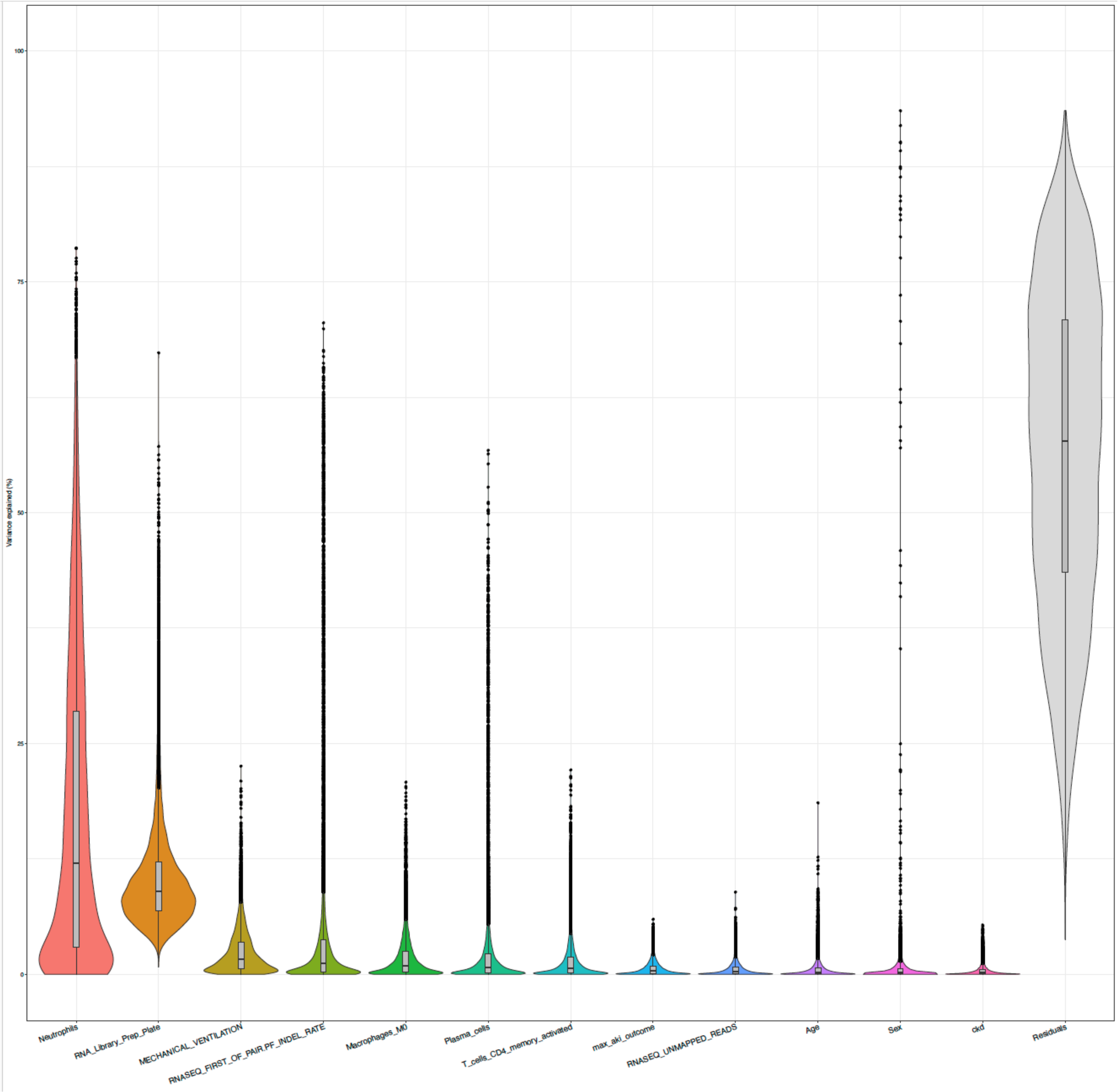

#### Differential expression analysis:

The volcano plot **(Fig 2B Main Manuscript)** displayed an unequal dispersion (in the context of number of genes and the extent of fold change) across the up and downregulated genes. The dispersion around the downregulated gene expressions shows fold change as low as 0.3 (70% lower expression than controls), while the upregulated genes showed dispersions with the highest fold change of 2 (100% higher expression than controls). Of the differentially expressed genes, we found signatures of known markers of AKI and renal tubular injury, regulatory genes that might cause activation of other signaling mechanisms, and genes known to be involved in sepsis associated AKI.

The top canonical pathways with more than 50% significantly downregulated genes included eiF2/eif4 signaling, oxidative phosphorylation, mitochondrial dysfunction mTOR signaling and NF-kB signaling. Canonical pathways with more than 50% significantly downregulated genes included eiF2/eif4 signaling, oxidative phosphorylation, mitochondrial dysfunction, *mTOR* signaling, Th17 activation, and NF-kB signaling **(Supplementary Fig S2A)**. In contrast, the *HOTAIR* regulatory pathway, estrogen receptor signaling pathway, and *PPARa/RXRa* activation had more than 50% of the genes upregulated.

**Functional analysis of differential expression**

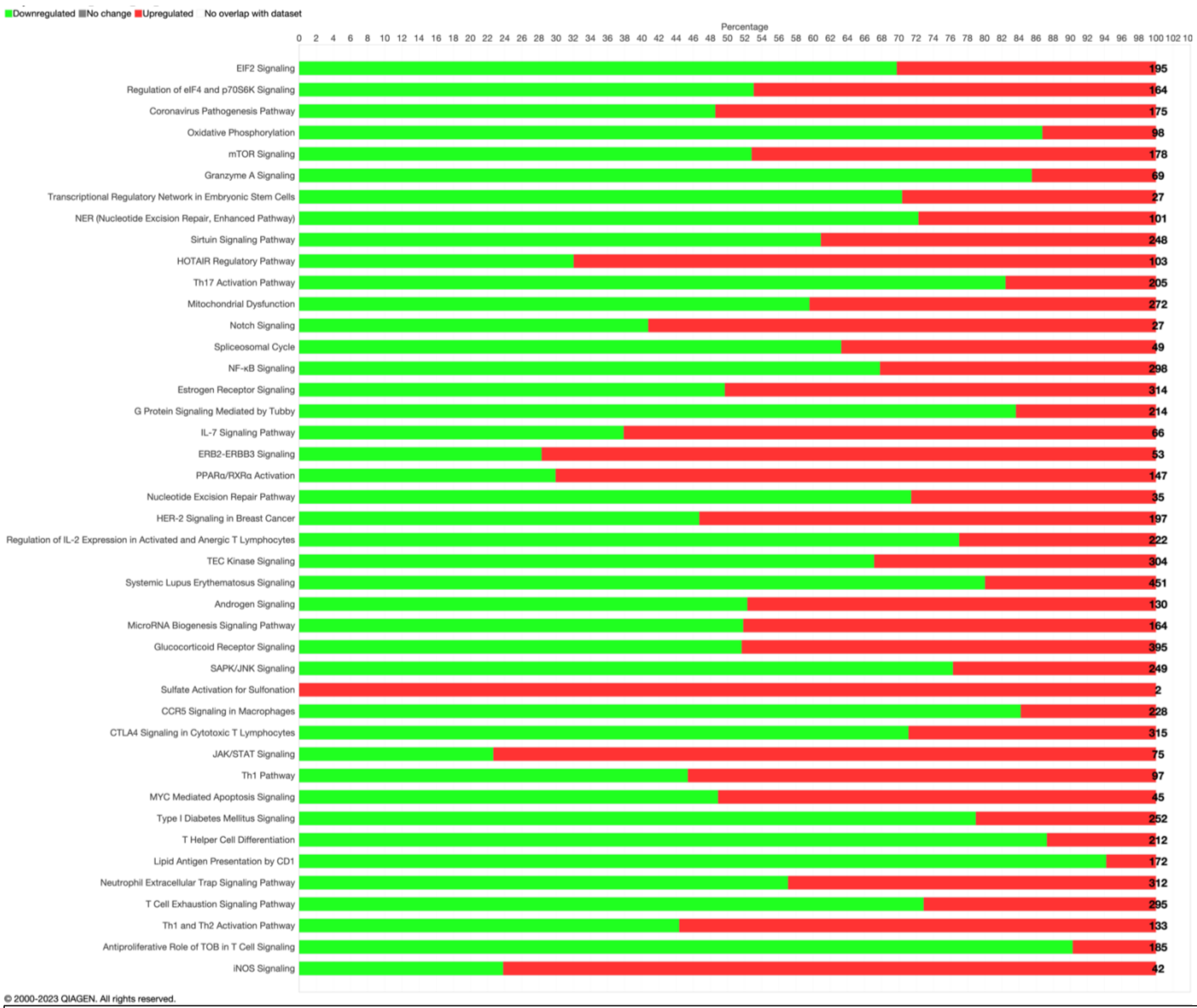

**Fig S2.A.** Canonical Pathways depicted by the percentage of up or downregulated genes in the pathway.

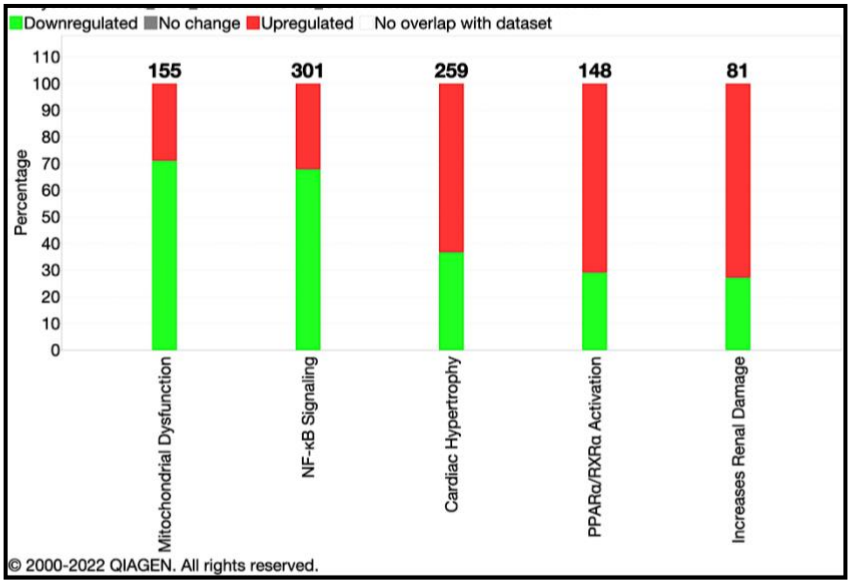

**Fig S2.B.** Curated pathways highlighting clinical pathology end points. Top clinical toxicities include 'mitochondrial dysfunction', 'NF-KB signaling' and 'cardiac hypertrophy', 'PPARa/RXRa activation' and an 'increase in renal damage'.

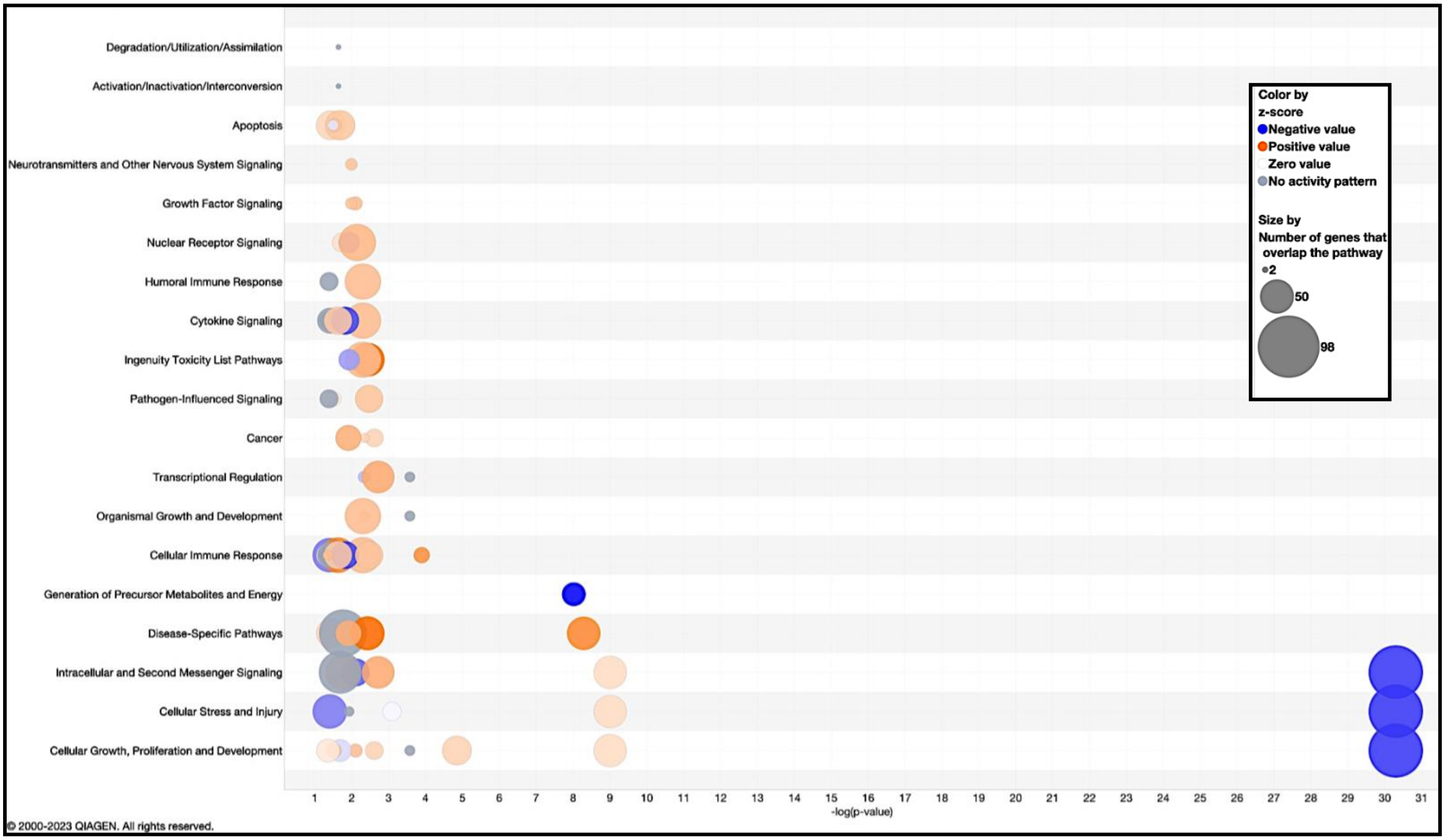

**Fig S2.C.** Bubble Chart showing the multiple pathway categories that the canonical pathways belong to. Chief among the major categories of cellular processes to be affected were pathways involved in "cellular oxidative stress response", "inflammation", "cytokine signaling" and "apoptosis".

##### Comparison with Sepsis driven AKI (s-AKI)

The study was designed to identify multi-omic changes (metabolomic, proteomic and transcriptomic) changes in patients with Systemic Inflammatory Response Syndrome (SIRS) who developed sepsis. SIRS Patients (n=125) who had sepsis and developed AKI were tagged as cases(n=64) and the rest were controls (n=61). To validate the similarity between the molecular signatures of COVID-associated AKI and sepsis-associated AKI, we performed unpaired student's T-Test between the differentially expressed list of significant genes in our cohort against the published signatures of sepsis-associated AKI. In addition, we also performed enrichment of the published canonical pathways in the sepsis associated AKI cohort within the identified significant canonical pathways in our cohort(see Main Manuscript Methods and Results)

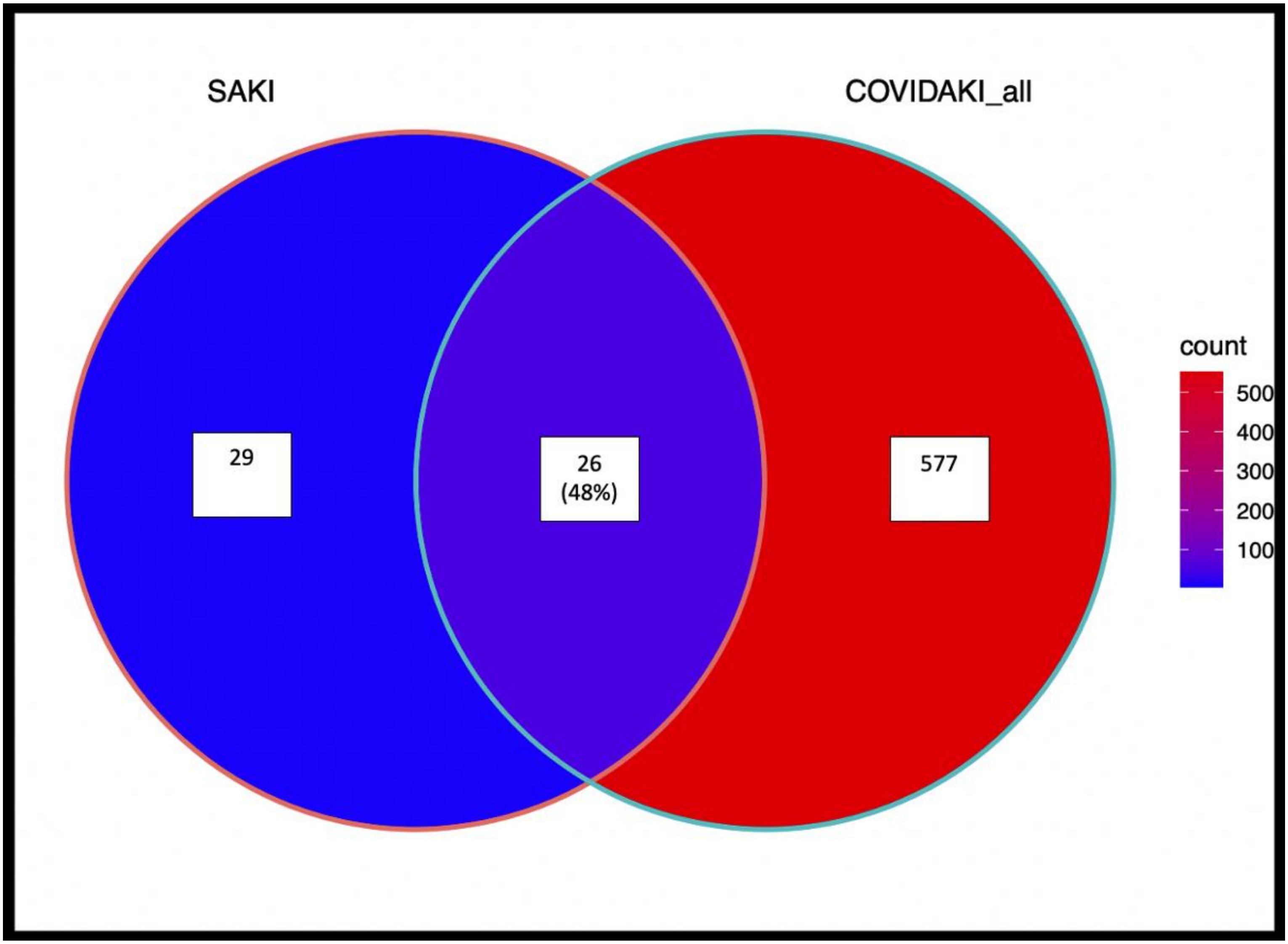

***Fig S4.A.*** *A Venn Diagram of the significant canonical pathways from a published s-AKI manuscript and obtained results from the c-AKI analyses reveals many of the s-AKI pathways are also involved in c-AKI.*

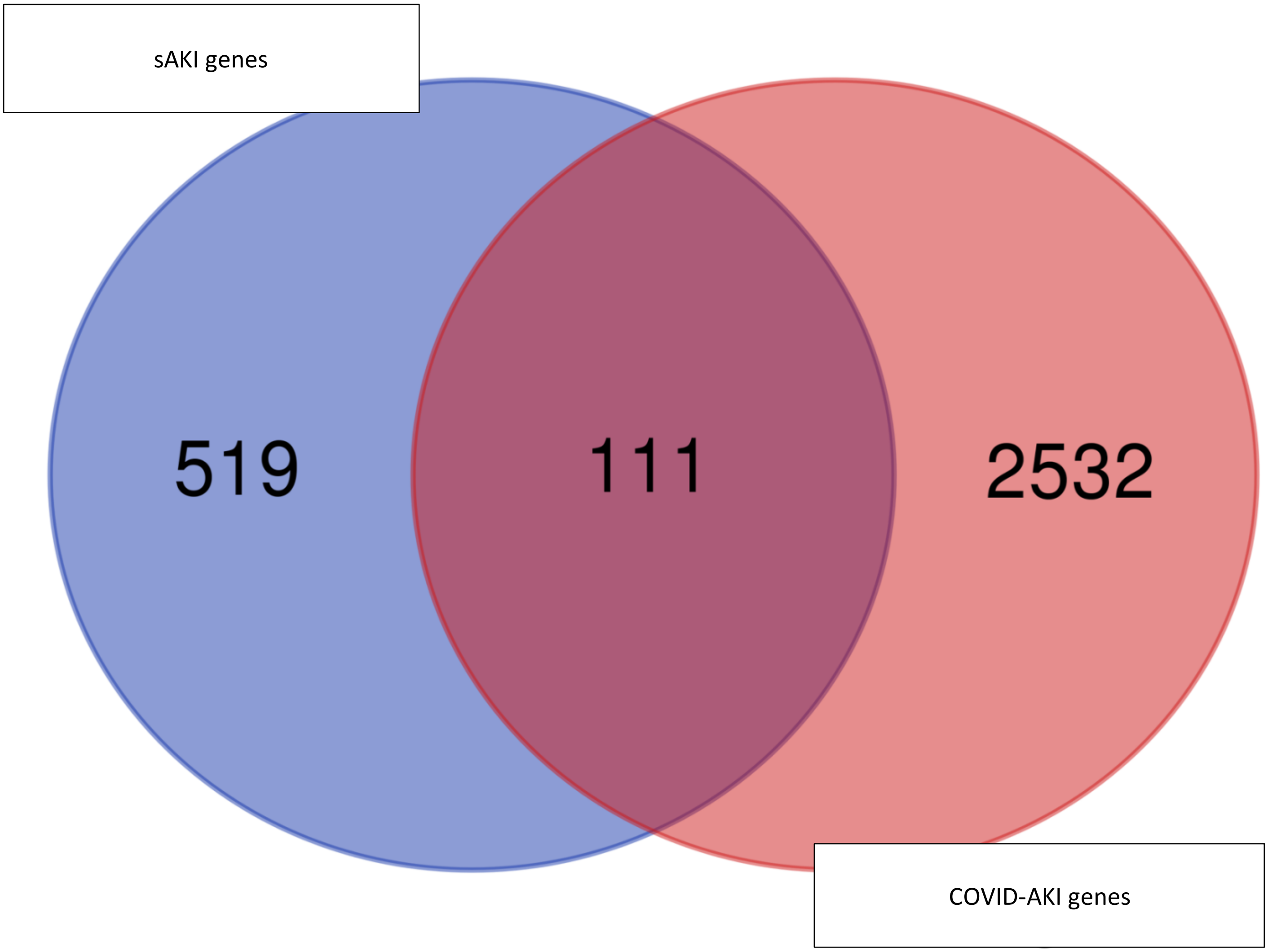

***Fig S4.B****. A Venn Diagram of the overlap of significant differentially expressed genes from a published s-AKI manuscript and obtained results from the c-AKI analyses reveals many of the s-AKI genes are also involved in c-AKI. 111 of the 630 sepsis genes were enriched in the COVID-AKI dataset as well.*

The Fisher exact test statistic value for enrichment of cAKI genes in sAKI cohort is $< 0.0001$. The result is significant at p-value $< 0.00001$.

#### Analysis of long-term kidney dysfunction from post-discharge eGFR measurements

We transformed the data using the *loess* smoothing function as implemented in the *ggplot2* R package.

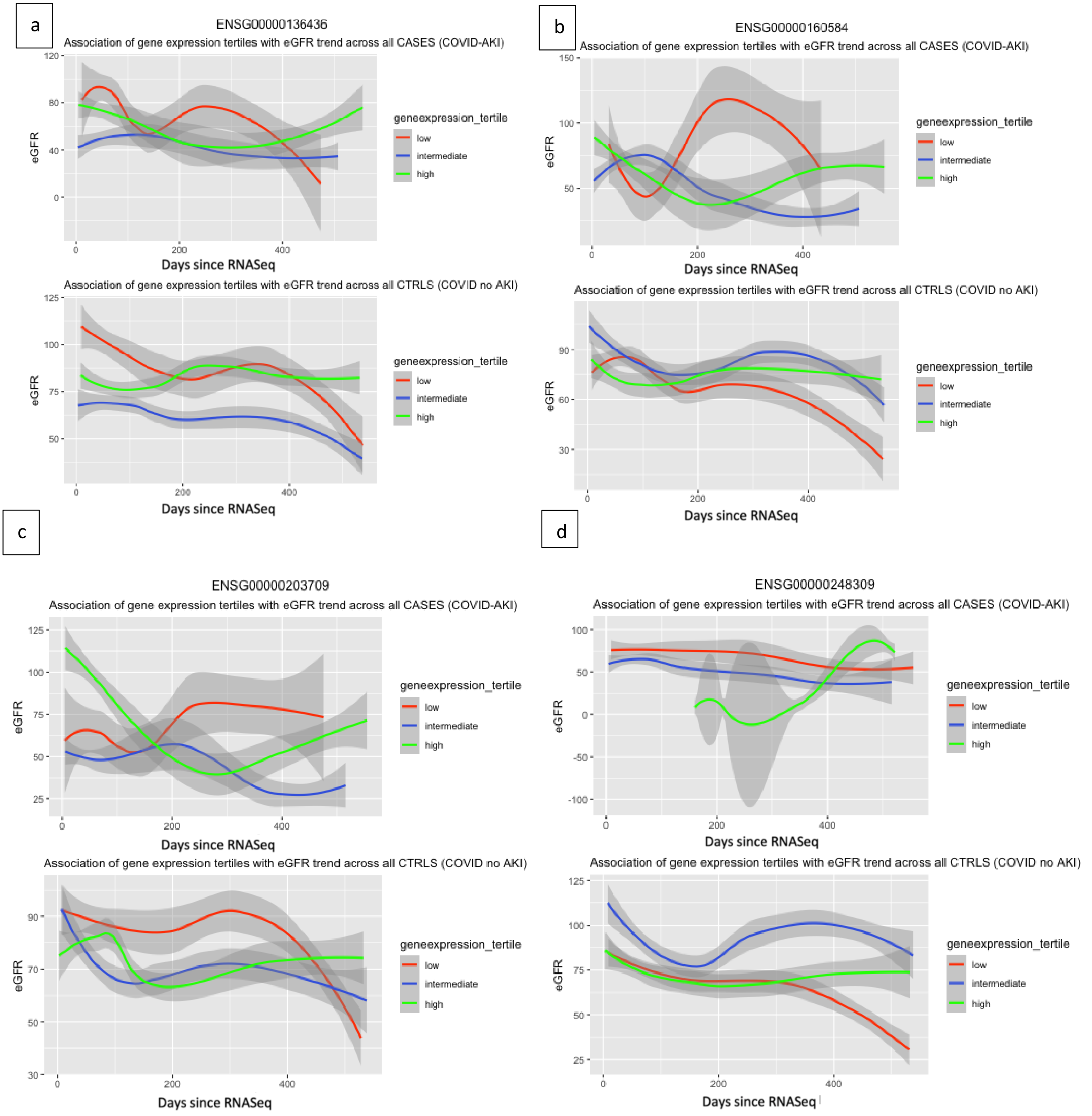

Fig **S5**. The figures above indicate the corelation of the top 3 up and down-regulated gene expression tertiles with eGFR trend across all cases (hospitalized COVID patients who developed AKI) and controls (hospitalized COVID patients who did not develop AKI). **S5.a.** ENSG00000136436 or CALCOCO2. **S5.b.** ENSG00000160584 or SIK3. **S5.c.** ENSG00000203709 or mir-29B. **S5.d.** ENSG00000248309 or MEF2C. **S5.e.** ENSG00000155666 or KDM8.

e

f

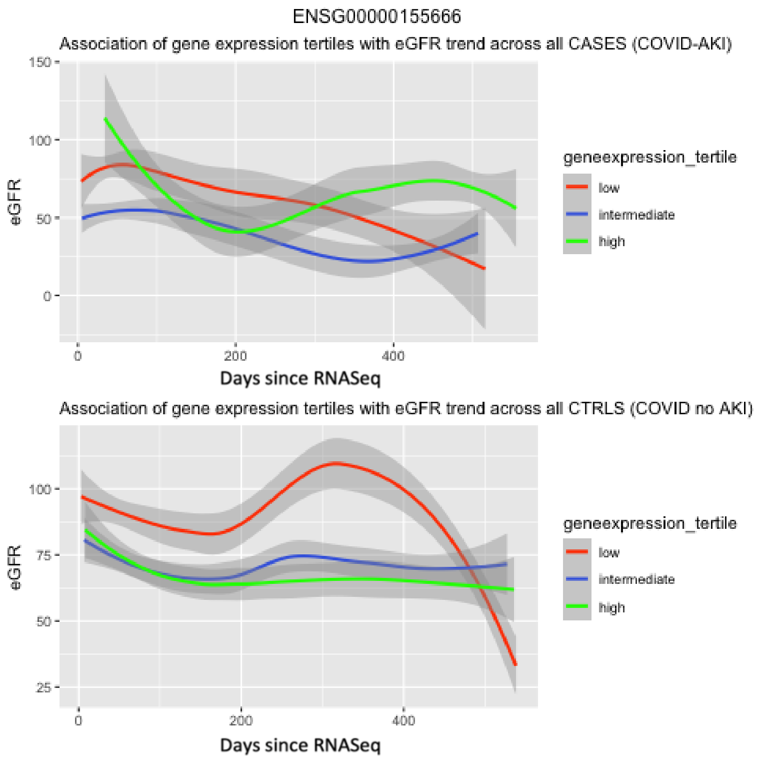

ENSG00000155666
Association of gene expression tertiles with eGFR trend across all CASES (COVID-AKI)
eGFR
150
100
50
0
0
200
400
Days since RNASeq
geneexpression_tertile
low
intermediate
high
Association of gene expression tertiles with eGFR trend across all CTRLS (COVID no AKI)
eGFR
100
75
50
25
0
200
400
Days since RNASeq
geneexpression_tertile
low
intermediate
high
